# Incubation Period and Reproduction Number for novel coronavirus (COVID-19) infections in India

**DOI:** 10.1101/2020.06.27.20141424

**Authors:** SR Patrikar, A Kotwal, VK Bhatti, A Banerjee, K Chatterjee, R Kunte, M Tambe

**Affiliations:** Armed Forces Medical College, Pune, India; Armed Forces Medical Services, India; Dr DY Patil Medical College, Pune, India; BJ Medical College, Pune, India

## Abstract

Novel coronavirus (COVID-19) rapidly spread from China to other parts of the world. Knowledge of incubation period and reproduction number is important in controlling any epidemic. The distribution of these parameters helps estimate the epidemic size and transmission potential of the disease. We estimated incubation period and reproduction number of COVID-19 for India utilizing data reported by Ministry of Health and Family Welfare (MoHFW), Government of India (GOI) and data in public domain. The mean incubation period seems to be larger at 6.93 (SD=±5.87, 95% CI: 6.11-7.75). and 95^th^ percentile estimate for best fit normal distribution is 17.8 days. Weibull distribution, the best fit for the reproduction number estimated pre lockdown reproduction number as 2.6 (95% CI=2.34 - 2.86) and post lockdown reduced to 1.57 (95% CI=1.3 – 1.84) implying effectiveness of the epidemic response strategies. The herd immunity is estimated between 36-61% for R_0_ of 1.57 and 2.6 respectively.

## Introduction

While the novel coronavirus (COVID-19) spread rapidly from China to other developed countries, India saw a steady flow of patients mod early March and by 10 May 2020, it had gripped the country with 63,420 confirmed cases and 2109 deaths^1-2^. The novel coronavirus (SARS-CoV-2) though related is distinct from severe acute respiratory syndrome (SARS) coronavirus and Middle East respiratory syndrome (MERS) coronavirus^3^. Many researchers struggled to estimate the magnitude of the epidemic wherein the epidemiological parameters remained uncertain. Knowledge of key epidemiological parameters including incubation period and reproduction number is important in controlling any epidemic. The distribution of these parameters helps estimate the epidemic size and transmission potential^3-7^ of the disease.

The incubation period is defined as the time from infection to the onset of illness^8^ and is crucial for epidemiological modelling in predicting the transmission dynamics, infectiousness and quarantine period^9^. It is also important for several important public health activities like length of active monitoring, surveillance and control.

The reproduction number (R_0_) is the most fundamental parameter in infectious disease dynamics describing the contagiousness or transmissibility of infectious agents and is defined as the average number of secondary cases caused by a single infectious individual in a entirely susceptible population^10^. An outbreak is expected to continue if R_0_ has a value >1 and to end if R_0_ is <1. R_0_ fluctuates if the rate of human–to-human or human and vector interactions varies over time or space. There exists scant evidence which supports the applicability of R_0_ outside the region for which the value was calculated^11^. Estimation of changes in transmission over time can provide insights into the epidemiological situation and identify whether outbreak control measures are adequate and are having the desired measurable effect, and help in undertaking midcourse corrections.

Herd immunity is defined as the resistance to the spread of a contagious disease within a population that results if a sufficiently high proportion of individuals are immune to the disease, especially through vaccination or immunity post natural infection^12^. When a high proportion of the population is immune, it is difficult for infectious diseases to spread, because there are not many people who can be infected and the transmission chain gets broken.

Our current understanding of these epidemiological parameters for India is limited. Hence this study was undertaken to address above issue and estimate incubation period and reproduction number of COVID-19 for India utilizing data reported by Ministry of Health and Family Welfare (MoHFW), Government of India (GOI) and the data available in public domain.

## Materials and Methods

### Data sources

The analysis is based on publicly available data. Data were retrieved from the official website of the MoHFW, GOI^1^ and covid19indi tracker^2^. The other sources for data aggregation and triangulation were, World Health Organization (WHO)^13^, Centre for Disease Control (CDC)^14^, update reports of Indian Council of Medical Research (ICMR)^15^, and India Situation Reports on Novel Coronavirus (2019-nCoV)^16^. Since the analysis is based on publicly available data ethics approval was not required.

Information on demographic characteristics, date on exposure and date of confirmation of disease status/illness onset were extracted for estimating incubation period and adjusted for delay-time. The delay time in getting the test results/diagnosis as confirmed case of COVID-19 was adjusted based on the delay time available from states and MoHFW reports. For estimating reproduction number, details of the source of infection from positive case giving number of probable cases infected by suspect was extracted. The number of people who contracted the disease from primary case.

### Statistical distributions for Incubation period and reproduction number

The incubation period data post adjustment of delay-time in test results was subjected to best fit model. Besides normal distribution four other commonly used incubation period distribution (Weibull, Log normal, Gamma and Erlang) were fitted. Estimation of the median incubation period, mean (sd), and quantiles (5^th^, 25^th^, 75^th^, 95^th^ and 97^th^) was also done.

For reproduction number, the best fit model (Weibull distribution) was used. Besides the best, fit three more distributions based on review of literature for distribution of reproduction number (lognormal, Gamma and Exponential) were considered. The values of R_0_ were estimated for pre and post lockdown period to evaluate the impact of the epidemic control measures instituted by the Government of India. The herd immunity (HI) estimate was based on R_0_ value^12^. (HI= (R_0_ – 1) or R_0_ = 1 − 1/R_0_) The statistical software used were IBM SPSS Statistics for Windows version 23.0 (SPSS Inc., Chicago, Ill., USA) and EASY-FIT, a software system for data fitting in dynamical systems.

## Results

Data on 268 lab confirmed cases were extracted, the range of age was from 1.5 years to 89 years with a mean age of 36.45 years (SD=±17.27). Ratio of male to female was 1.5:1 with 60.3% males and 39.7% females.

### Incubation Period

Using date of exposure and date of confirmation of disease status/illness onset, the estimates of incubation period were determined. Table 1 gives the incubation period estimates for various distributions. We fitted five distributions to the data: Normal, Weibull, Log normal, Gamma and Erlang. The normal distribution provided best fit for data with median and mean incubation period of 6.93 (SD=±5.87, 95% confidence interval CI: 6.11-7.75). The incubation period ranged from 1 to 19.26 days (5^th^ percentile to 97^th^ percentile) for this best fit. The Weibull distribution was the best next fit with mean incubation period to be higher than normal distribution with mean of 8.17 (SD=8.10). Figure 1 show cumulative distribution functions for best fit. The median incubation period for other distributions ranged 3.46 to 6.06. The probability and cumulative distribution functions for various distributions in order of best fit is provided in the supplementary.

**Table 1:** Incubation Period distribution.

| Incubation Period** | Normal | Weibull | Log Normal | Gamma | Erlang |
| --- | --- | --- | --- | --- | --- |
| Mean | 6.93 | 8.17 | 8.72 | 7.64 | 4.99 |
| SD | 5.87 | 8.10 | 13.69 | 6.17 | 4.9 |
| 95% CI for Mean | 6.11-7.75 | 7.04-9.3 | 6.82-10.6 | 6.78-8.5 | 4.32-5.68 |
| 5<sup>th</sup> percentile | 1 | 0.4 | 0.75 | 0.93 | 0.25 |
| 25<sup>th</sup> percentile | 2.96 | 2.36 | 2.21 | 3.13 | 1.43 |
| **Median (50 percentile)** | **6.93** | **5.68** | **4.69** | **6.06** | **3.46** |
| 75 percentile | 11.81 | 11.34 | 9.95 | 10.46 | 6.92 |
| 95 percentile | 17.81 | 22.42 | 29.32 | 19.78 | 14.95 |
| 97 percentile | 19.26 | 26.57 | 33.14 | 22.6 | 17.5 |

\*\*Adjusted for delay time in test results

**Figure 1:**
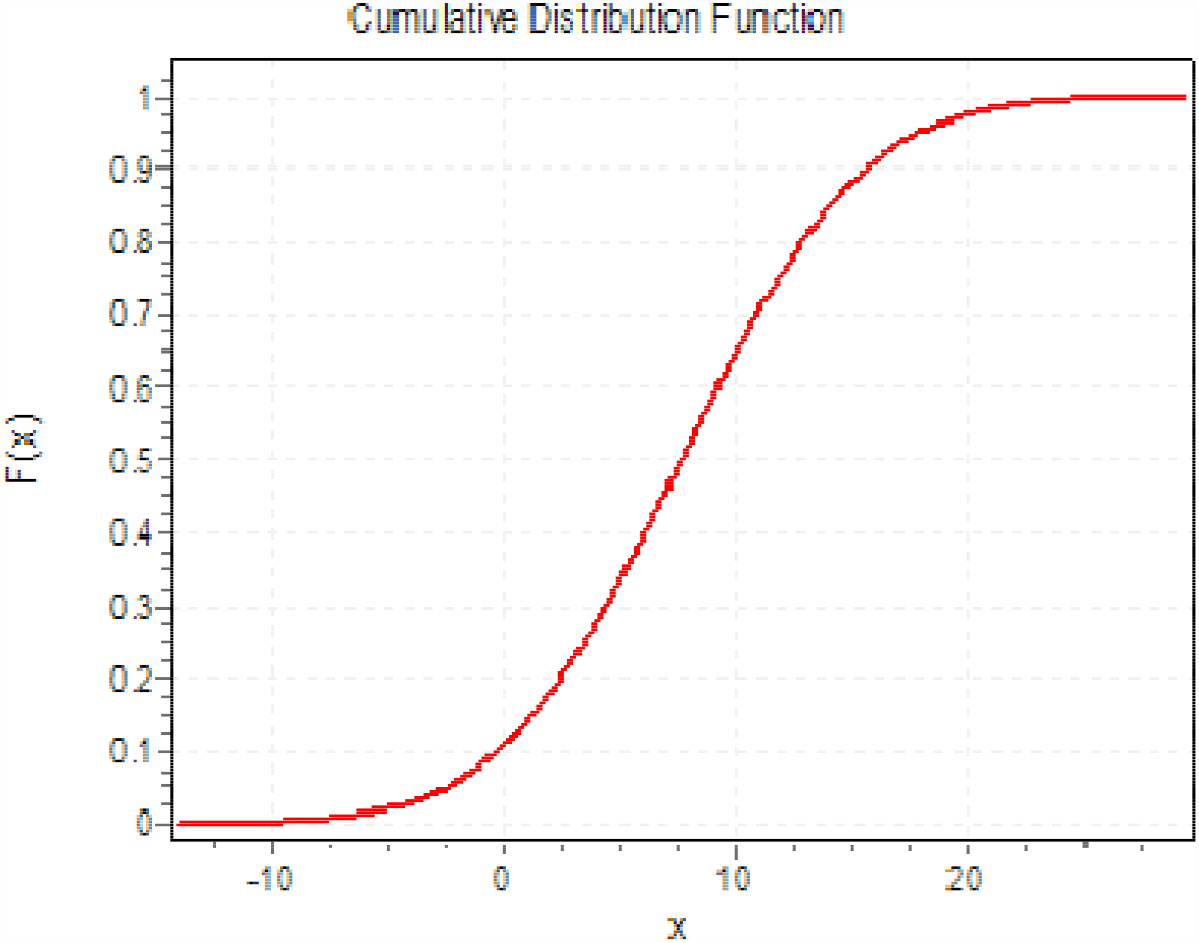
Cumulative distribution function for best fit Normal distribution for Incubation period for India

### Reproduction Number and Herd Immunity

The reproduction number was estimated for days before lockdown and post lockdown to assess the impact of various control measures to include social distancing adopted by India. Weibull distribution was the best fit for the reproduction number followed by Log Normal and Gamma distribution. We estimated the initial reproduction number before lockdown by GOI to be 2.6 (95% CI=2.34 - 2.86) and post lockdown the reproduction number reduced to 1.57 (95% CI=1.3 – 1.84). The herd immunity is estimated in the range of 61-62%. Table 2 gives the descriptive statistics of reproduction number before and post lock down.

**Table 2:** Reproduction Number and Herd Immunity.

| **Reproduction number before lock down** |  |  |  |  |
| --- | --- | --- | --- | --- |
|  | Weibull | Log Normal | Gamma | Exponential |
| Mean | 2.6 | 2.5 | 2.61 | 2.63 |
| SD | 1.89 | 2.1 | 2.46 | 2.6 |
| Median | 2.19 | 1.91 | 1.87 | 1.8 |
| 95% Confidence interval | 2.34-2.86 | 2.21-2.79 | 2.27-2.95 | 2.27-2.99 |
| Herd Immunity | 61.5% | 61% | 61.7% | 62% |
| **Reproduction number post lock down** |  |  |  |  |
|  | Weibull | Log Normal | Gamma | Exponential |
| Mean | 1.57 | 1.55 | 1.57 | 1.59 |
| SD | 0.87 | 0.81 | 1.03 | 1.50 |
| Median | 1.46 | 1.37 | 1.35 | 1.10 |
| 95% Confidence interval | 1.3-1.84 | 1.3-1.8 | 1.25-1.89 | 1.12-2.06 |
| Herd Immunity | 36% | 35% | 36% | 37% |

## Discussion

This data driven paper to the best of our knowledge presents the estimates of incubation period and reproduction number of COVID-19 in India for the first time. We characterised the distribution of incubation period and reproduction number for COVID-19 for India. We estimated that the incubation period follows the best fit of normal distribution with around 7 days ranging from 1 to 22 day with 95^th^ percentile of the distribution at 17.8 days. The incubation period seems to be longer for India as compared to 2 to 10 days by WHO^13^ and 2-12 days by ECDC^17^. The United States’ CDC^18^ estimates the incubation period for COVID-19 to be between 2 and 14 days. Most of existing studies are on data originating from Wuhan, China. A study on travellers from Wuhan on 88 cases estimated the mean incubation period to be 6.4 days with a range from 2.1 to 11.1 days. The upper limit of 11.1 days was considered conservative^19^. Two other studies from Wuhan, China on early transmission dynamics of COVID-19 estimated the mean incubation period as 5.2 days (95% CI, 4.1 to 7.0), with the 95th percentile of the distribution at 12.5 days^20^ and other study with 2-14 days with Log Normal as best fit^21^. A study by John Hopkins Bloomberg School of Public Health on regions and countries outside Hubei^22^ gave median incubation period as 5.1 days (95% CI, 4.5 to 5.8 days), and also stated that 97.5% of those who develop symptoms will do so within 11.5 days (CI, 8.2 to 15.6 days) of infection. Understanding incubation period is crucial so as to introduce and implement effective quarantine duration and preventing the spread of the virus.

Weibull distribution seems to be the best fit for reproduction number distribution in India. The reproduction number for India is estimated to be 2.6 in pre lock down phase and reduced to around 1.6 post lockdown phase with a herd immunity threshold of 61.5%. Though the value of R_0_ has reduced from 2.6 in pre lock down period to 1.57 in post lock down period. i.e. 60.38% reduction it is still greater than 1. COVID-19 epidemic will increase as long as R_0_ is greater than 1 and control efforts to bring R_0_ below 1 needs to be implemented aggressively.

A report based on the impact of the interventions across 11 European countries estimated a posterior mean of 0.97 [0.14-2.14] for Norway and 2.64 [1.40-4.18] for Sweden, with an average of 1.43 across the 11 country posterior means, a 64% reduction compared to the pre-intervention values^23^. A study from Wuhan, Hubei Province, China^20^ estimated an R_0_ of approximately 2.2, however this estimate was limited to early epidemic period up to January 4. The median R0 from Diamond Princess cruise ship^24^ was estimated to be 2.28 (95% CI=2.06-2.52). Switzerland^25^ estimated the basic reproduction number R_0_ of COVID-19 at 2.78 (95% confidence interval, CI: 2.51 - 3.11). Transmission decreased with the strengthening of social distancing measures by 89% (95% CI: 83%-94%) which brought the R_0_ to below 1 at 0.32. Study^25^ by Wu et. al. estimated the reproductive number for COVID-19 as 2·68 (95% CI: 2·47–2·86). The wide range of R_0_ from different studies indicates the challenges in estimating R_0_. Additionally, R_0_ is dynamic in the sense that any factor having the potential to influence the contact rate, including population density, social distancing, and seasonality will ultimately affect R_0_.

Our estimate of herd immunity threshold as 61% (with R_0_ =2.6) is staggering as 826,873,494 people need to be infected to achieve herd immunity in India which may result in high death rate. Many countries have implemented aggressive lock downs however some countries like Sweden had a different approach to tackle COVID-19 by taking individual responsibility for social distancing and keeping society functioning with no official lock down by the Sweden government and still successful in keeping the reproduction number in control^26^. Long term lockdowns are not sustainable as its ill effects impacts many other health programmes. As per WHO^27^ the weekly detection of new tuberculosis cases in India has gone down by nearly 75 per cent during the COVID-19 lockdown. The malaria modelling analysis by the Global Malaria Programme at WHO^28^ and the HIV modelling study, convened by the WHO and UNAID^29^, predicts the collateral damage of COVID will be as devastating as pandemic, with millions succumbing to preventable, treatable illness and disease. However, in absence of vaccine and other uncertainties regarding the virus, herd immunity is debatable issue as a preventive measure and achieving herd immunity is likely to be a long-drawn process.

### Limitations of the study

The data on complete information for estimating incubation period as well reproduction number could be extracted for limited number of cases. Also, the time delay of test results, with varying turn around time among States, for estimating incubation period was averaged for India based on the public domain information and media reports.

## Conclusions

This study provides the estimates for key epidemiological charachteristics of COVID-19 in India. The mean incubation period seems to be larger at 6.93 and 95^th^ percentile estimate for best fit normal distribution to be 17.8 days. Best fit for reproduction number follows weibull distribution. Our estimates for pre lockdown reproduction number was 2.6. and post lockdown the reproduction number reduced to 1.57. This implies and shows that the epidemic response strategies adopted by India are effective. However the herd immunity is estimated in the range of 36-61% for R_0_ of 1.57 and 2.6 respectively.

## Data Availability

The analysis is based on publicly available data

https://www.mohfw.gov.in/

https://www.covid19india.org/

## References

1. Home | Ministry of Health and Family Welfare | GOI [Internet]. Available from: https://www.mohfw.gov.in/

2. COVID19INDIA tracker. COVID-19-India: Patient Database [Accessed on 19 April 2020]. https://www.covid19india.org/.

3. Zhu N, Zhang D, Wang W, Li X, Yang B, Song J, et al. A novel coronavirus from patients with pneumonia in China, 2019. N Engl J Med. 24 Jan 2020: NEJMoa2001017. https://doi.org/10.1056/NEJMoa2001017 PMID: 319789453

4. Imai N, Dorigatti I, Cori A, Donnelly C, Riley S, Ferguson NM. Report 2: Estimating the potential total number of novel Coronavirus cases in Wuhan City, China. London; 2020. Available from: https://www.imperial.ac.uk/media/imperialcollege/medicine/sph/ide/gida-fellowships/2019-nCoVoutbreak-report-22-01-2020.pdf

5. Leung K, Wu JT, Leung GM. Nowcasting and forecasting the potential domestic and international spread of the 2019-nCoV outbreak originating in Wuhan, China: a modelling study. Lancet. 31 Jan 2020. Available from: https://www.thelancet.com/journals/lancet/article/PIIS0140-6736(20)30260-9/fulltext

6. Nishiura H, Jung SM, Linton NM, Kinoshita R, Yang Y, Hayashi K, et al. The Extent of Transmission of Novel Coronavirus in Wuhan, China, 2020. J Clin Med. 2020;9(2):330. https://doi.org/10.3390/jcm9020330 PMID: 31991628

7. Imai N, Cori A, Dorigatti I, Baguelin M, Donnelly CA, Riley S, et al. Report 3: Transmissibility of 2019-nCoV. London; 2020. Available from: https://www.imperial.ac.uk/media/imperialcollege/medicine/sph/ide/gida-fellowships/Imperial-2019nCoV-transmissibility.pdf

8. Zhao S, Lin Q, Ran J, Musa SS, Yang G, Wang W, et al. Preliminary estimation of the basic reproduction number of novel coronavirus (2019-nCoV) in China, from 2019 to 2020: A data-driven analysis in the early phase of the outbreak. Int J Infect Dis. 30 Jan 2020. Available from: https://www.sciencedirect.com/science/article/pii/S1201971220300539

9. Linton NM, Kobayashi T, Yang Y, et al. Incubation period and other epidemiological characteristics of 2019 novel coronavirus infections with right truncation: a statistical analysis of publicly available case data. J Clin Med. 2020;9. [PMID: 32079150] doi:10.3390/jcm9020538

10. Stephen AL, Kyra HG, Qifang Bi MHS. Forrest KJ, Qulu Zheng, Hannah RM, Andrew SA, Nicholas GR, Justin Lessler, The Incubation Period of Coronavirus Disease 2019 (COVID-19) From Publicly Reported Confirmed Cases: Estimation and Application, Annals of Internal Medicine, Vol. 172 No. 9, May 2020

11. Delamater PL, Street EJ, Leslie TF, Yang YT, Jacobsen KH. Complexity of the basic reproduction number (R0). Emerg Infect Dis. 2019 Jan. https://doi.org/10.3201/eid2501.171901

12. Paul Fine, Ken Eames, David L. Heymann, “Herd Immunity”: A Rough Guide, Clinical Infectious Diseases, Volume 52, Issue 7, 1 April 2011, Pages 911–916, https://doi.org/10.1093/cid/cir007

13. WHO Coronavirus disease (COVID-19) Pandemic, [Accessed on 21 April 2020] Available from https://www.who.int/emergencies/diseases/novel-coronavirus-2019

14. Centers of Disease Control and prevention, CDC, https://www.cdc.gov/coronavirus/2019-nCov/index.html

15. Indian Council of Medical Research, ICMR [Accessed on 21 April 2020] https://icmr.nic.in/content/covid-19

16. Novel Coronavirus (2019-nCoV) Situation Report-7 - World Health Organization (WHO), January 27, 2020

17. European Centre for Disease Prevention and Control (ECDC). Q & A on novel coronavirus. Stockholm: ECDC; 2020. [Accessed 21 April 2020]. Available from: https://www.ecdc.europa.eu/en/novel-coronavirus-china/questions-answers

18. Symptoms of Novel Coronavirus (2019-nCoV) – CDC

19. Backer Jantien A, Klinkenberg Don, Wallinga Jacco. Incubation period of 2019 novel coronavirus (2019-nCoV) infections among travellers from Wuhan, China, 20–28 January 2020. Euro Surveill. 2020;25(5):pii=2000062. https://doi.org/10.2807/1560-7917.ES.2020.25.5.2000062

20. Li Q, Guan X, Wu P, et al. Early transmission dynamics in Wuhan, China, of novel coronavirus-infected pneumonia. N Engl J Med. 2020. [PMID: 31995857] doi:10.1056/NEJMoa2001316

21. Linton NM, Kobayashi T, Yang Y, et al. Incubation period and other epidemiological characteristics of 2019 novel coronavirus infections with right truncation: a statistical analysis of publicly available case data. J Clin Med. 2020;9. [PMID: 32079150] doi:10.3390/jcm9020538

22. Lauer SA, Grantz KH, Bi Q, et al. The Incubation Period of Coronavirus Disease 2019 (COVID-19) From Publicly Reported Confirmed Cases: Estimation and Application. Ann Intern Med. 2020; [Epub ahead of print 10 March 2020]. doi: https://doi.org/10.7326/M20-0504

23. Seth Flaxman Swapnil Mishra, Axel Gandy, H Juliette T Unwin, Helen Coupland, Thomas A Mellanet. Al., Report 13 - Estimating the number of infections and the impact of non-pharmaceutical interventions on COVID-19 in 11 European countries, Imperial College COVID-19 Response Team, 30 March 2020

24. Sheng Zhanga, MengYuan Diaob, Wenbo Yuc, Lei Peic, Zhaofen Lind, Dechang Chena, Estimation of the reproductive number of novel coronavirus (COVID-19) and the probable outbreak size on the Diamond Princess cruise ship: A data-driven analysis, International Journal of Infectious Diseases 93 (2020) 201–204

25. Christian L. Althaus, Real-time modeling and projections of the COVID-19 epidemic in Switzerland, Institute of Social and Preventive Medicine, University of Bern, Switzerland 24 April 2020 [Accessed on 12 May 2020], https://ispmbern.github.io/covid-19/swiss-epidemic-model/

25. Wu TJ, Leung K, Leung GM, Nowcasting and forecasting the potential domestic and international spread of the 2019-nCoV outbreak originating in Wuhan, China: a modelling study, The Lancet, Volume 395, Issue 10225, 29 February–6 March 2020, Pages e4

26. Sweden: estimate of the effective reproduction number 29 april 2020, https://www.folkhalsomyndigheten.se/contentassets/4b4dd8c7e15d48d2be744248794d1438/sweden-estimate-of-the-effective-reproduction-number.pdf

27. Developed by Stop TB Partnership in collaboration with Imperial College, Avenir Health, Johns Hopkins University and USAID.The Potential Impact Of The Covid-19 Response On Tuberculosis In High-Burden Countries:A Modelling Analysis [Accessed on 14 May 2020], http://www.stoptb.org/assets/documents/news/Modeling%20Report_1%20May%202020_FINAL.pdf

28. World Health Organization (WHO), The potential impact of health service disruptions on the burden of malaria: a modelling analysis for countries in sub-Saharan Africa, Global Malaria Programme

29. L. Jewell Britta; Mudimu, Edinah; Stover, John; Kelly, Sherrie L.; Phillips, Andrew (2020): Potential effects of disruption to HIV programmes in sub-Saharan Africa caused by COVID-19: results from multiple mathematical models. figshare. Preprint. https://doi.org/10.6084/m9.figshare.12279914.v1

